# Validation of the GCS-Pupil scale in Traumatic Brain Injury Incremental prognostic performance of pupillary reactivity with GCS in the prospective observational cohorts CENTER-TBI and TRACK-TBI

**DOI:** 10.1101/2024.06.05.24308424

**Authors:** Rick J.G. Vreeburg, Florian D. van Leeuwen, Geoffrey T. Manley, John K. Yue, Paul M. Brennan, Xiaoying Sun, Sonia Jain, Thomas A. van Essen, Wilco C. Peul, Andrew I.R. Maas, David K. Menon, Ewout W. Steyerberg, the CENTER-TBI TBI TRACK-TBI participants and investigators and the members of the clinical working group of the NIH-NINDS initiative on classification and nomenclature of TBI

## Abstract

**Objective:** To compare the incremental prognostic value of pupillary reactivity as captured in the GCS-Pupils score (GCS-P) or added as separate variable to the Glasgow Coma Scale (GCS) in traumatic brain injury (TBI).

**Methods:** We analyzed patients enrolled between 2014 and 2018 in the Collaborative European NeuroTrauma Effectiveness Research in Traumatic Brain Injury (CENTER-TBI, n=3521) and the Transforming Research and Clinical Knowledge in Traumatic Brain Injury (TRACK-TBI, n=1439) cohorts. We used logistic regression to quantify the prognostic performances of GCS-P versus GCS according to Nagelkerke’s R^2^. Endpoints were mortality and unfavorable outcome (Glasgow Outcome Scale-Extended score 1-4) at 6 months after injury. We estimated 95% confidence intervals with bootstrap resampling to summarize the improvement in prognostic capability.

**Results:** GCS as a linear score had a R^2^ of 24% (95% confidence interval [CI] 17-30) and 30% (95%CI 17-43) for mortality and 29% (95%CI 25-34) and 38% (95%CI 29-47) for unfavorable outcome in CENTER-TBI and TRACK-TBI respectively. In the meta-analysis, pupillary reactivity as a separate variable improved the R^2^ by an absolute value of 6% and 2% for mortality and unfavorable outcome (95%CI 4.0-7.7 and 1.2-3.0, respectively), with half the improvement captured in the GCS-P score (3%, 95%CI 2.1-3.3 and 1%, 95%CI 1-1.7, respectively).

**Conclusions:** GCS-P has a stronger association with outcome after TBI than the GCS alone. However, for prognostic models, inclusion of GCS and pupillary reactivity as separate scores is preferable.

## Background

Traumatic brain injury (TBI) is a pressing, multifaceted health concern affecting millions of people worldwide annually.^1–3^ The initial clinical severity of TBI is commonly reported according to the Glasgow Coma Scale (GCS), which is often trichotomized into three classes of severity: mild (GCS 13-15), moderate (GCS 9-12) and severe (GCS≤8). This tripartite division is embedded in clinical practice and research, but neglects patient experiences and the relevant heterogeneity within the divisions. Consequently, therapeutic nihilism may result in patients with presumed “severe” injuries, while disabling complaints and symptoms may be disregarded in patients with presumed “mild” injuries.^4,5,6^ The NIH-NINDS has implemented an international initiative to develop a novel approach to TBI classification, culminating in a workshop in Bethesda, USA in January 2024.^7^

In preparation of this workshop, the working group on clinical assessment considered the relative value of using the full GCS or the GCS-Pupils score (GCS-P) for classifying the clinical severity of TBI. The GCS-P was proposed by Brennan and colleagues in 2018.^8,9^ In this novel scale one point is deducted from the GCS score for each unreactive pupil, resulting in a score ranging from 1 to 15, which intuitively has merit. Both the GCS and pupillary reactivity serve as important features in surgical decision-making, and lower scores are associated with poorer outcome. Moreover, the GCS (or its motor component), and pupillary reactivity are important components of well validated multifactorial prognostic models, such as the International Mission for Prognosis and Analysis of Clinical Trials in TBI (IMPACT) and Corticosteroid Randomization after Significant Head Injury (CRASH) models.^10–12^

With the simplicity of the GCS-P being comparable to the GCS and the information yield potentially greater, it shows potential to better characterize TBI. Studies conducted on the IMPACT and CRASH datasets have shown that merging of GCS and pupillary reactivity into the GCS-P may have comparable information yield compared to using GCS and pupillary reactivity as separate factors, but these were mainly focused on patients with moderate to severe TBI, and it is uncertain if this may hold across all severities.^8,9^ Uncertainty further exists if a summary score like the GCS-P provides similar prognostic information as the inclusion of these features separately in a prognostic model. We aimed to analyze the association of GCS and GCS-P with outcome in large contemporaneous datasets including TBI of all severities and to explore their prognostic performance relative to each other and to a model including GCS and pupillary reactivity as separate prognostic factors.

## Methods

### Study population

This study used data from 2 large multicenter, prospective, observational cohorts: the Collaborative European NeuroTrauma Effectiveness Research in TBI (CENTER-TBI) and the Transforming Research and Clinical Knowledge in Traumatic Brain Injury (TRACK-TBI) studies.^13,14^ CENTER-TBI included TBI patients between 2014 and 2017 presenting to one of 65 participating centers across Europe. TRACK-TBI enrolled TBI patients presenting to the emergency department between 2014 and 2019 from 18 United States (US) level 1 trauma centers through convenience sampling. CENTER-TBI and TRACK-TBI are registered on ClinicalTrials.gov (number NCT02210221 and NCT02119182 respectively).

All adults (≥ 18 years) with a TBI recruited to these two studies were included in the current analysis. Exclusion criteria were: (1) missing baseline motor score or pupillary reactivity scorings or (2) missing Glasgow Outcome Scale-Extended (GOS-E) scores.

### GCS(-P) & Pupillary reactivity

For CENTER-TBI, baseline GCS and pupillary reactivity were defined, using IMPACT methodology as the most recent not missing value between emergency room (ER) discharge (post-stabilisation) and pre-hospital assessment. Untestable eye (swelling) and verbal (intubation) components of the GCS were imputed with the number “1”.^15^ In TRACK-TBI, the baseline GCS and pupillary reactivity were defined as the assessment at ER presentation. Missing eye and verbal components were imputed as follows:

In case only verbal score is untestable: Total GCS = 0.55+1.45*[eye]+1.44*[mot].

In case both Eye and verbal score is untestable: Total GCS = 0.6+2.4*[mot].

The GCS was analyzed as an ordinal scale starting at 3 (lowest score) and ending at 15 (highest score). Pupillary reactivity was expressed using the Pupil Reactivity Score (PRS) which is scored based on 3 levels, namely both reacting, one reacting and none reacting, resulting in a score of 0, −1 or −2. The GCS and PRS were combined into an integrated GCS-P score, according to predefined methodology in the initial GCS-P study, subtracting the PRS from the GCS, yielding scores from 1 to 15.^8^

### Outcomes

Patient (long-term) functional outcome was expressed using the GOS-E scale at 6 months after injury. GOS-E is an extended measure used to quantify functional outcome and consists of an 8-point scale ranging from 1 (death) to 8 (upper good recovery).^16^ The categories 2 (vegetative state) and 3 (lower severe disability) were merged as these could not be differentiated on assessments performed by postal questionnaire. In CENTER-TBI, missing outcomes at 6 months after injury were imputed using a multinomial model if assessments at one or more other time points were available. In TRACK-TBI, only patients with available GOSE scores at 6 months were included. The primary outcomes are unfavorable outcome, which was defined as a GOS-E score lower than “lower moderate disability” (<5) and mortality, defined as patient death within 6 months after injury.

### Statistical analysis

Logistic regression modelling was used to analyze the relationships between GCS, PRS, GCS-P and patient outcome (GOS-E and mortality). Nagelkerke’s pseudo R^2^ was used as primary measure to quantify the prognostic capability of the included parameters.^17^ Nagelkerke’s R^2^ is calculated at the log likelihood scale. It is a measure of how much better the model fits the data compared to a model with no predictors. Nagelkerke’s R^2^ can be interpreted as a measure of the proportion of variation explained in the dependent variable (the six-month outcome) that is explained by the independent variables (predictors) in a logistic regression model.^18,19^ The uncertainty of the R^2^ estimate was quantified by bootstrap resampling (5000 repetitions). We also estimated the increase in R^2^, the 1′R^2^, for GCS-P versus GCS models and GCS plus PRS versus GCS-P models within each bootstrap sample. The distribution of bootstrapped R^2^ estimates was used to estimate 95% confidence intervals (CI) for R^2^ and differences between R^2^ estimates. A pooled estimate across the CENTER-TBI and TRACK-TBI studies was estimated with inverse variance weighting. The regression analyses yielded estimations of odds ratios (OR) and 95% CI. The OR indicates the odds of unfavorable outcome (over favorable outcome) per 1-point increase in the GCS, PRS or GCS-P scales.

A subgroup analysis was performed to assess the prognostic capabilities (mortality and unfavorable outcome) of GCS and GCS-P versus a model with both GCS and PRS in patients with moderate to severe TBI (GCS 3-12). Moreover, an additional age-stratified subgroup analysis was performed on 3 different age groups based on the age distribution in the cohorts, namely age <45, age 45-64, age ≥65 years.

Statistical analysis was performed using R version 4.1.2.^20^

## Results

### Demographics and baseline characteristics

The total CENTER-TBI and TRACK-TBI cohorts consisted of 4509 and 2552 patients respectively. Of those patients, 3521 from CENTER-TBI and 1439 from TRACK-TBI were eligible for primary analysis (Figure 1).

**Figure 1.**
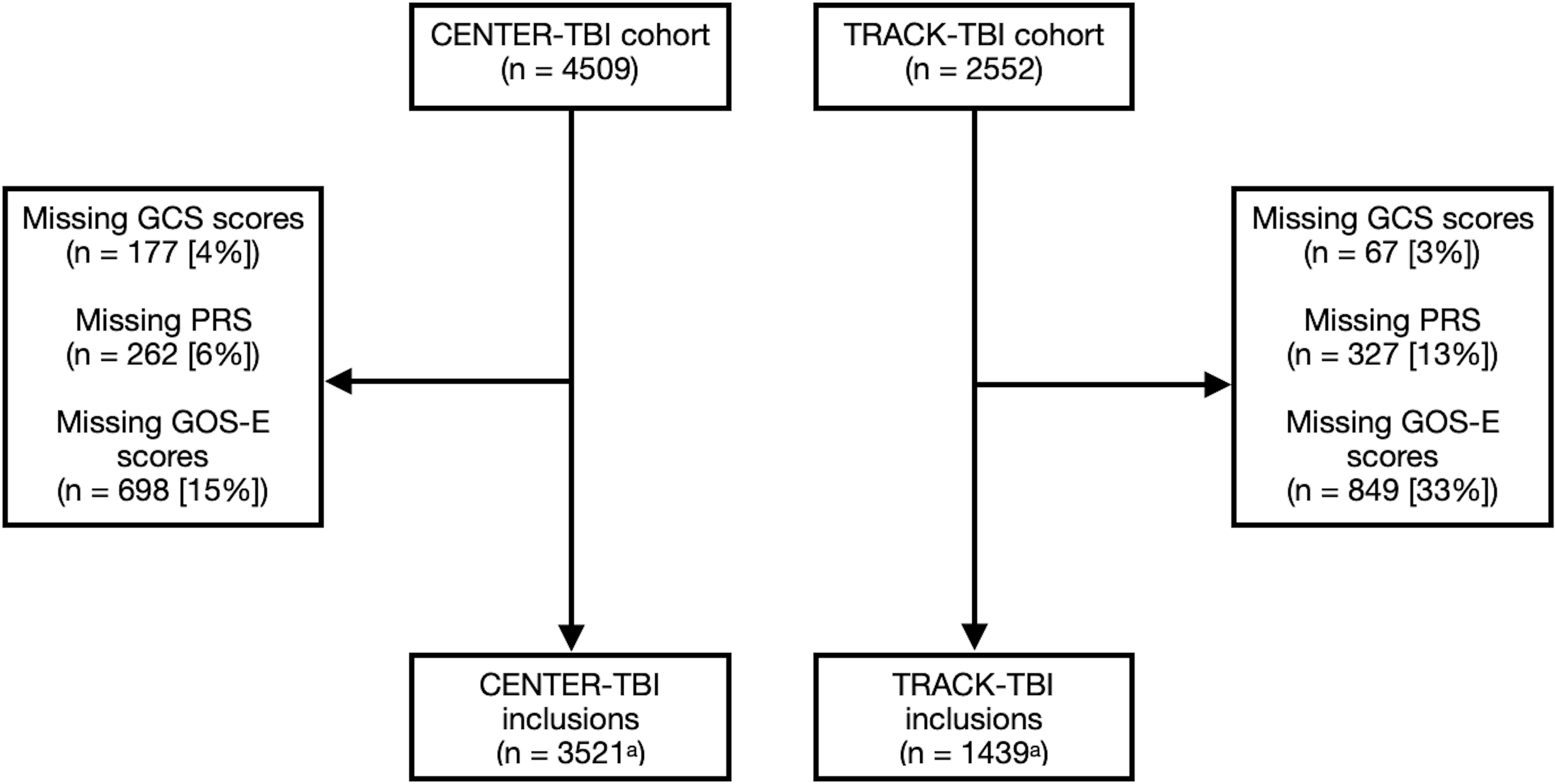

The included patients from both studies were similar in sex, median GCS, PRS and CT imaging variables such as the presence of epidural hematoma (EDH), acute subdural hematoma (ASDH), traumatic subarachnoid hemorrhage (tSAH) and diffuse axonal injury (DAI) (Table 1). TRACK-TBI patients were generally younger than CENTER-TBI patients (median age: 39 years vs 51 years, respectively). Furthermore, mild TBI was more frequent in TRACK-TBI (78%) patients compared to CENTER-TBI (67%). Moreover, cerebral contusions (36% and 25%, respectively) and midline shift (16% and 11%, respectively) were more prevalent in CENTER-TBI compared to TRACK-TBI respectively.

**Table 1.** | Baseline patient characteristics.

|  | CENTER-TBI | TRACK-TBI |
| --- | --- | --- |
| **No of patients** | 3521 | 1439 |
| **Age (median [IQR])** | 51 [30, 67] | 39 [26, 56] |
| **Male sex (%)** | 2341 (67) | 982 (68) |
| **GCS scores (median [IQR])** | 15 [9, 15] | 15 [13, 15] |
| **TBI severity** |  |  |
| Mild (GCS 13-15) | 2360 (67) | 1122 (78) |
| Moderate (GCS 9-12) | 316 (9) | 70 (5) |
| Severe (GCS < 9) | 845 (24) | 247 (17) |
| **Pupils (%)** |  |  |
| Both reacting | 3141 (89) | 1333 (93) |
| One reacting | 137 (4) | 31 (2) |
| Both unreacting | 243 (7) | 76 (5) |
| **Epidural hematoma (%)** | 382 (11) | 122 (9) |
| **Subdural hematoma, acute (%)** | 1107 (33) | 431 (31) |
| **Cerebral contusion (%)** | 1219 (36) | 341 (25) |
| **Traumatic subarachnoid haemorrhage (%)** | 1092 (35) | 536 (39) |
| **Midline shift<sup>b</sup> (%)** | 540 (16) | 154 (11) |
| **Diffuse axonal injury (%)** | 341 (11) | 132 (10) |
| **6-month unfavorable outcome (GOS-E < 5)** | 878 (25) | 200 (14) |
| **6-month mortality**<br>**(GOS-E = 1)** | 418 (12) | 98 (7) |

*Abbreviations:* CENTER-TBI, Collaborative European NeuroTrauma Effectiveness Research in Traumatic Brain Injury; GOS-E, Glasgow Outcome Scale Extended; IQR, interquartile range; No, number; TRACK-TBI, Transforming Research and Clinical Knowledge in Traumatic Brain Injury

### Distribution of outcomes

Mortality and unfavorable outcome were more frequent in CENTER-TBI (12% and 25%) versus TRACK-TBI (7% and 14%). However, there were no large differences in the distribution of unfavorable outcome and mortality per GCS and PRS and within GCS-P scores (Supplemental Table 1 & 2). In CENTER-TBI, a small decrease in mortality at both GCS and GCS-P = 7 and 8 was observed while mortality increased again at GCS and GCS-P = 9 (Figure 2). Moreover, low GCS-P (1-2) displayed higher percentages of mortality and unfavorable outcome compared to low GCS (3-4). Mild and moderate TBI displayed similar distributions (Figure 2). In TRACK-TBI, low GCS-P (1-2) displayed lower mortality percentages at 6 months compared to CENTER-TBI (Figure 2 and Supplemental Table 3).

**Figure 2.**
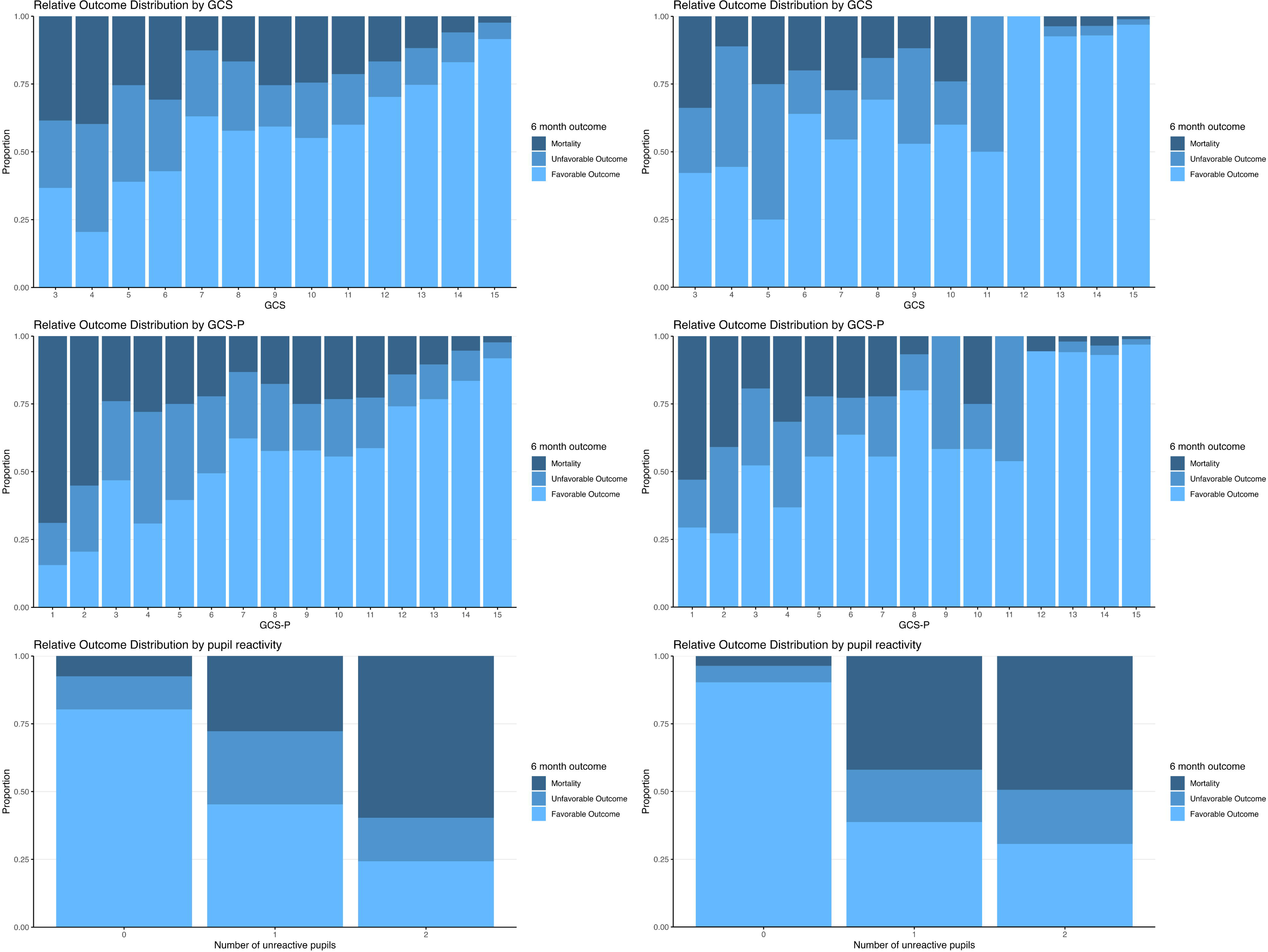

### Associations of GCS(-P) and PRS with outcome

The explained variance was higher for the regression model containing GCS and PRS (R^2^ 30% and 35%, Table 2) as separate predictors compared to the model containing GCS-P alone (R^2^ 27% and 33%) for mortality in both CENTER-TBI and TRACK-TBI respectively (Figure 3).

**Figure 3.**
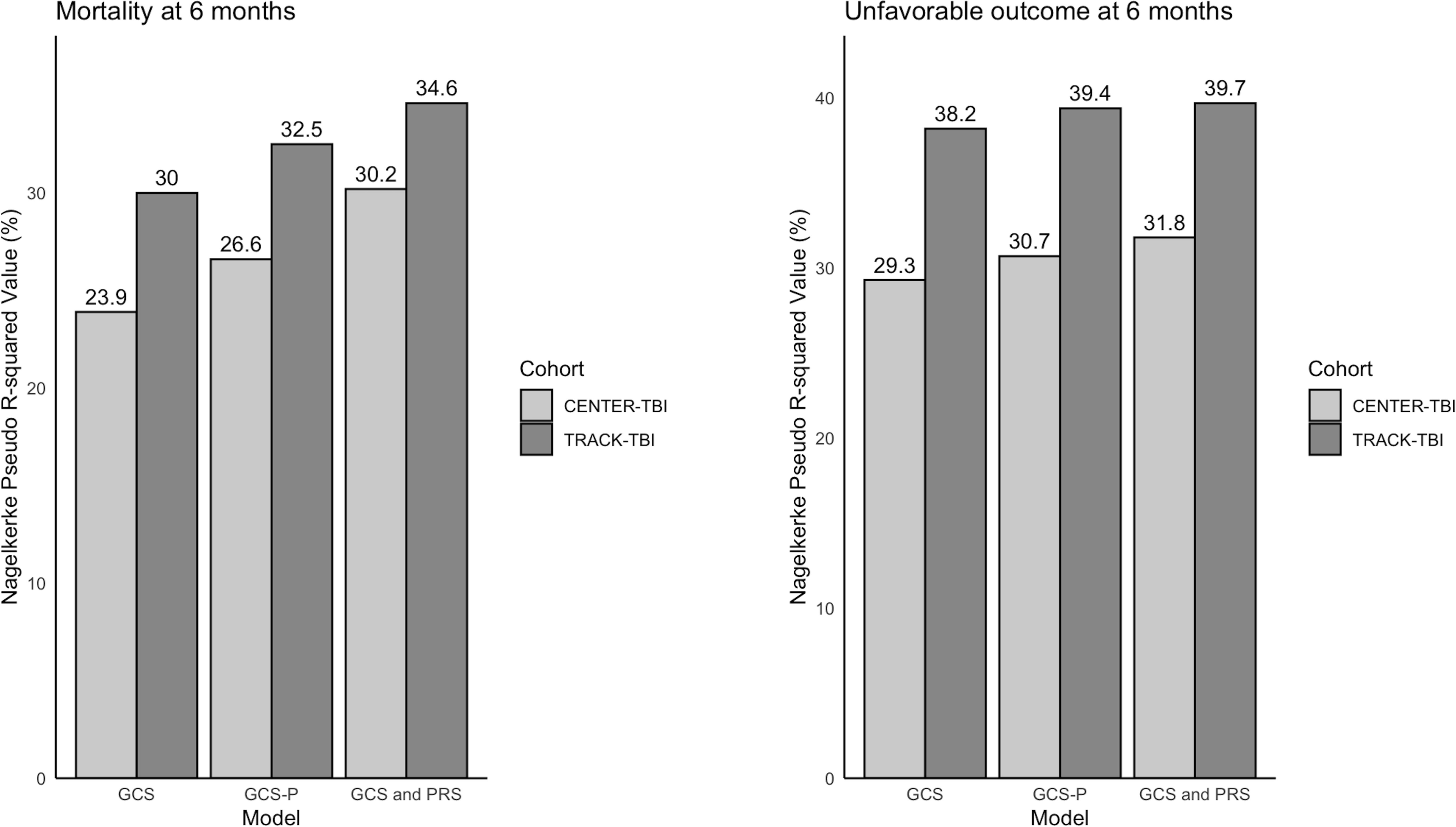

**Table 2.**
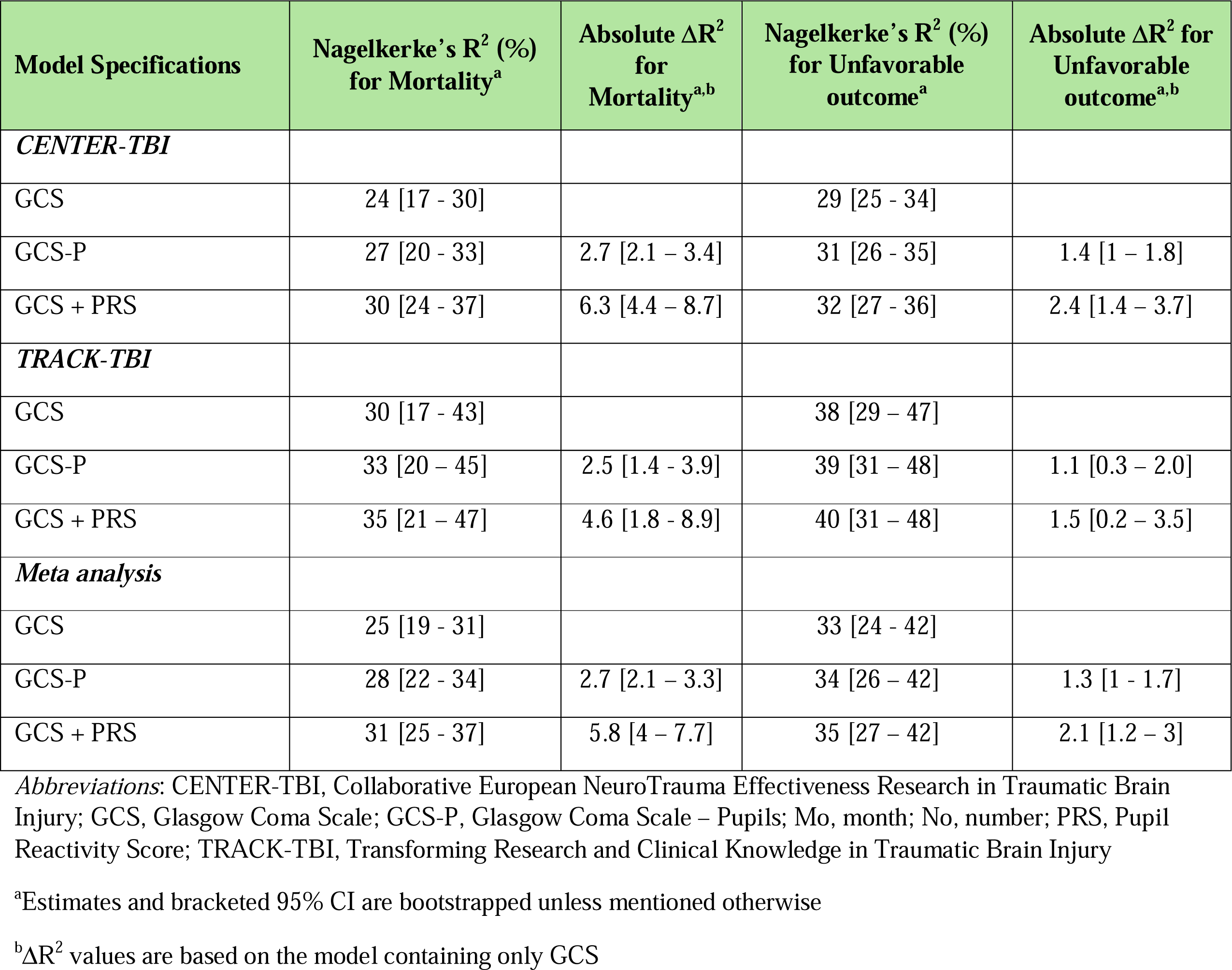
| Overall predictive performance of different regression models.

A model containing only GCS had the lowest model performance in CENTER-TBI and TRACK-TBI (R^2^ 24% and 30%, respectively). GCS had a lower range of predicted risk compared to GCS-P and GCS together with PRS (Supplemental Figure 1). Regarding unfavorable outcome, explained variance was highest in the model containing GCS and PRS as separate predictors compared to GCS-P in both CENTER-TBI and TRACK-TBI (GCS and PRS: R^2^ 32% and 40% & GCS-P: R^2^ 31% and 39%, respectively, Table 2). Moreover, a model containing only GCS had the lowest model performance in both cohorts (R^2^ 29 % and 38% for CENTER-TBI and TRACK-TBI respectively). In a meta-analysis across studies, pupils as a separate variable improved the R^2^ by an absolute value of 6% and 2% for mortality and unfavorable outcome, with half the improvement captured in the GCS-P score (3% and 1%, respectively, Table 2, Figure 3 and 4).

**Figure 4.**
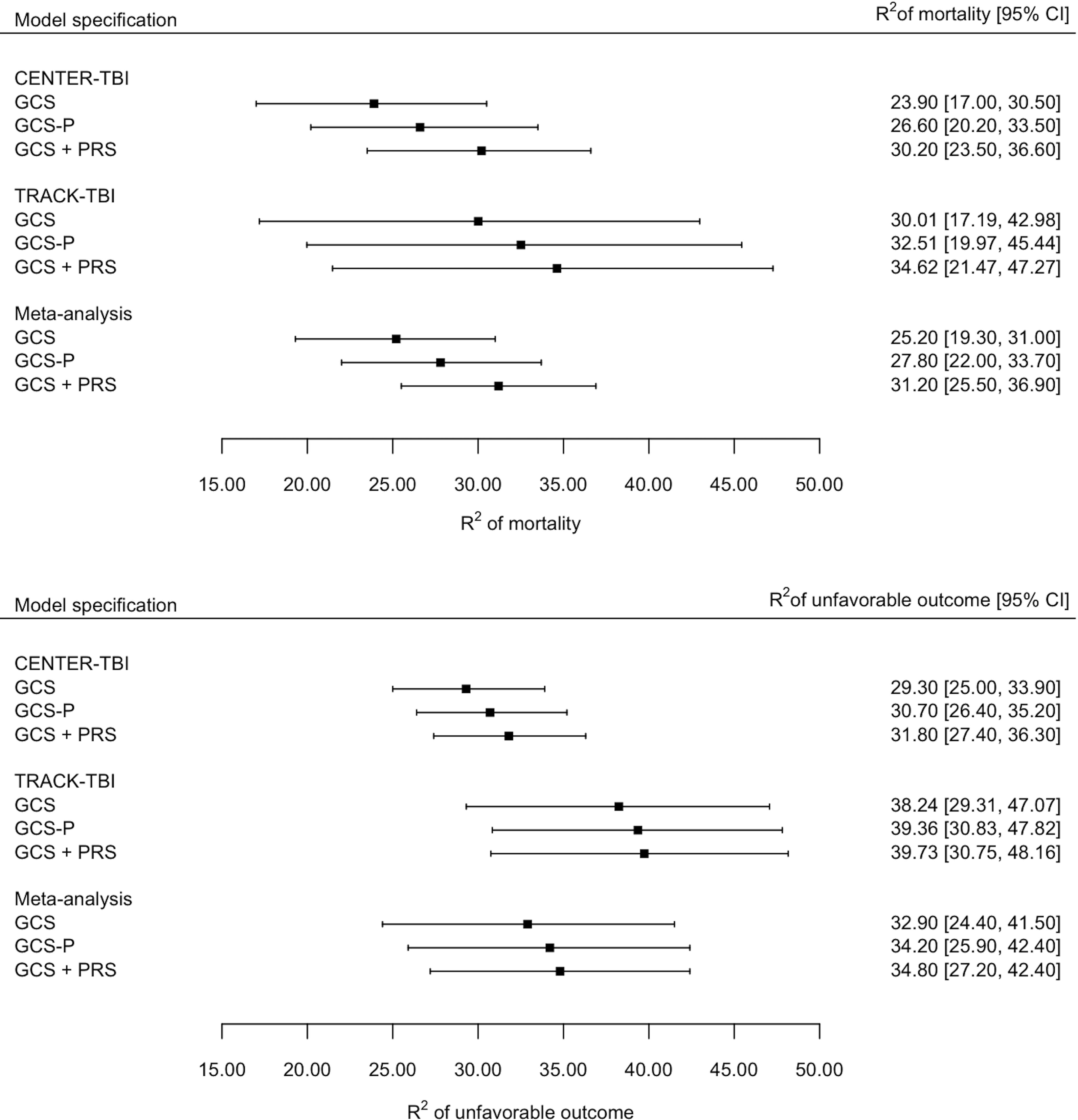

For both cohorts, in a logistic regression model containing GCS, an incremental 1-point increase in GCS significantly decreased the odds for mortality within 6 months after injury (OR 0.79, 95%CI 0.77-0.81 and OR 0.75, 95%CI 0.72-0.79 for CENTER-TBI and TRACK-TBI respectively, Table 3). Similarly, more favorable GCS-P showed comparable odds ratios in both CENTER-TBI and TRACK-TBI (OR 0.79, 95%CI 0.77-0.81 and OR 0.76, 95%CI 0.73-0.79, respectively). In a model with both GCS and PRS, incremental 1-point decreases in PRS decreased the odds of mortality more strongly (OR 0.4, 95%CI 0.33-0.45 and OR 0.43, 95% CI 0.32-0.58 for CENTER-TBI and TRACK-TBI, respectively) than the odds for GCS (OR 0.84, 95%CI 0.82-0.86 and 0.80, 95%CI 0.76-0.85).

**Table 3.** | Odds ratios & coefficients of independent baseline variables predicting outcome.

| Model Specifications | Odds ratio for Mortality<br>(95%CI interval) | Model Coefficient<br>for Mortality | Odds ratio for<br>Unfavorable outcome<br>(95%CI interval) | Model Coefficient<br>for Unfavorable<br>outcome |
| --- | --- | --- | --- | --- |
| ***CENTER-TBI*** |  |  |  |  |
| GCS | 0.79 (0.77 – 0.81) | -0.24 | 0.78 (0.77 – 0.8) | -0.24 |
| GCS-P | 0.79 (0.77 – 0.81) | -0.23 | 0.79 (0.78 – 0.8) | -0.24 |
| GCS + PRS<sup>a</sup> | 0.84 (0.82 – 0.86) | -0.18 | 0.81 (0.79 – 0.83) | -0.21 |
|  | & | & | & | & |
|  | 0.4 (0.33 – 0.45) | -0.92 | 0.5 (0.42 – 0.59) | -0.68 |
|  | respectively | respectively | respectively | respectively |
| ***TRACK-TBI*** |  |  |  |  |
| GCS | 0.75 (0.72 – 0.79) | -0.28 | 0.74 (0.71 – 0.77) | -0.3 |
| GCS-P | 0.76 (0.73 – 0.79) | -0.28 | 0.75 (0.73 – 0.78) | -0.29 |
| GCS + PRS<sup>a</sup> | 0.8 (0.76 – 0.85) | -0.22 | 0.77 (0.74 – 0.8) | -0.27 |
|  | & | & | & | & |
|  | 0.43 (0.32 – 0.58) | -0.84 | 0.56 (0.42 – 0.76) | -0.58 |
|  | respectively |  | respectively |  |

*Abbreviations:* CENTER-TBI, Collaborative European NeuroTrauma Effectiveness Research in Traumatic Brain Injury; CI, confidence interval; GCS, Glasgow Coma Scale; GCS-P, Glasgow Coma Scale – Pupils; IQR, interquartile range; Mo, month; No, number; PRS, Pupil Reactivity Score
<sup>a</sup>Odds ratios are expressed using decreases in PRS score, e.g. lower number of unreactive equally lower odds of mortality and unfavorable outcome

When predicting unfavorable outcome (GOS-E 1-4) using comparable models, similar associations were found (Table 3). Again, in a model with both GCS and PRS, a poorer PRS was more strongly associated with a lower odds of unfavorable outcome in both CENTER-TBI and TRACK-TBI patients (OR 0.5, 95%CI 0.42-0.59 and OR 0.56, 95%CI 0.42-0.76, respectively) than a lower GCS score (OR 0.81, 95%CI 0.-0. and OR 0.77, 95%CI 0.-0., respectively).

### Subgroup analysis

In the subgroup of 1161 patients in CENTER-TBI and 317 patients in TRACK-TBI with moderate and severe TBI (GCS 3-12), strong correlations were confirmed for both GCS-P, GCS and GCS and PRS with similar differences in explained variability (Supplemental Tables 4, Supplemental Figure 2). In the age-stratified subgroups, similar differences in explained variability between GCS-P, GCS and GCS and PRS were found, with greater explained variance in younger patients in CENTER-TBI and older patients in TRACK-TBI (Supplemental Table 4).

## Discussion

This study confirmed that in all models explored, pupillary reactivity adds important prognostic information over the GCS. However, the usage of PRS and GCS alone attributed to the greatest increment in 1′R^2^ for mortality and unfavorable outcome (5.8% and 2.1%), with half of this variance being captured by GCS-P (2.7% and 1.3%).

These results are largely in line with those reported in the original paper on GCS-P using the IMPACT and CRASH cohorts.^8^ In these studies, the addition of PRS to GCS as separate predictors in a regression model increased the 1′R^2^ by 4% and 3.2% with a 1′R^2^ of 3.4% and 2.2% between GCS and GCS-P for mortality and unfavorable outcome, respectively, leading the authors of the original paper to suggest that the GCS-P is a valuable summary score over the GCS alone. Both studies show an incremental advantage of the GCS and PRS as separate predictors across the broader spectrum of TBI included in the CENTER-TBI and TRACK-TBI studies. The replication of the seminal results of Brennan and colleagues in our study is remarkable in the presence of disparities between cohorts. A major difference is the fraction of mild TBI patients (GCS>8): 78% in CENTER-TBI and 67% in TRACK-TBI compared to 21% in the combined IMPACT and CRASH cohorts.^11^ Our primary analysis included all TBI patients with available GCS, PRS and GOSE, and thus were inclusive also of patients with GCS 15. Furthermore, patients in the original IMPACT and CRASH cohorts were considerably younger compared to CENTER-TBI and TRACK-TBI, with the median age in CENTER-TBI over 20 years higher than the median age in IMPACT.^11^ Pupillary abnormalities were also more common in the IMPACT cohort.

### Subgroup analyses

We performed a subgroup analysis on patients with moderate and severe TBI (GCS 3-12) to improve comparability between our study and that of Brennan and colleagues^9,10^. Comparable to the primary analysis, similar trends were observed in the prognostic information from GCS, PRS and GCS-P, but differences in explained variance between groups were more pronounced in moderate and severe TBI (Supplemental Table 4). The age-stratified subgroup analysis displayed variance between age groups regarding explained variability by the logistic regression models, but comparable trends and ΔR^2^ for the predictor variables in CENTER-TBI and TRACK-TBI

### Pros and cons of a summary score

Simplifications of the GCS inevitably convey less information and therefore loss of clinical value. Such considerations hold, for example, for the trichotomy of the GCS into mild, moderate and severe categories versus use of the full ordinal GCS sum score, but also to the GCS sum score compared to use of the underpinning eye, motor and verbal components, as their cumulative prognostic value is higher than that of the sum score alone.^21^

The integration of multiple clinical characteristics into a single score attempts to provide a single integrated articulation of TBI severity and status, without the loss of information. A careful consideration should be made whether any loss of information is acceptable in favor of utility and simplicity. A strength of the GCS-P is that it combines two of the most relevant clinical predictors for TBI into a summary score, and yet maintains the simplicity and ease of use of the GCS. Relative disadvantages include some loss of information compared to the use of GCS and PRS as separate predictors. Its utility as an overall parameter of injury severity is influenced by its “skewed” discrimination towards the high end of the severity spectrum. However, the subtraction of PRS from the GCS across the entire GCS range potentially reduces efficacy when non-reactive pupils occur at higher GCS scores. Therefore, GCS-P mainly provides additional value in patients with moderate to severe TBI which is in line with the index study by Brennan and Teasdale and the current study. Any decision about the use of an integrated score must also be balanced against potential implementation barriers, which may be substantial when introducing a modification of the GCS, which is deeply embedded in clinical practice.

Finally, different contributions from pupillary unreactivity and motor responses may add up to identical GCS-P scores but may have widely different clinical import and outcome. For example, a patient with a GCS of 3 and two reactive pupils (GCS-P = 3) may indeed have a very severe brain injury but could also have relatively less severe injury with examination confounded by alcohol, residual sedative drugs or a post-ictal state. However, a patient who has extensor motor responses and one unreactive pupil (E1 V1 M2 P-1) would be more uniformly likely to have a severe brain injury. Indeed, despite a small sample size and missing combinations, a comparison of outcomes of categories equaling a GCS-P score of 3 confirmed the variability in outcome in the first category, and a more dominant poor outcome in the second (Supplemental Table 2).

On balance, we consider the superiority of the GCS-P over the GCS modest, and from a prognostic perspective scoring of GCS and pupillary reactivity should be preferred. We further emphasize on a more general note that we would not recommend use of summary scores as replacements for the separate assessment of neurological status using GCS and PRS in individual patients.^8^

### Study strengths and weaknesses

The major strength of this study is its usage of prospectively collected data from 2 large multicenter observational studies across all severities in different continents with standardized data collection, thus ensuring generalizability of findings.

Limitations of our study include the restriction of data collection to North America and Europe, while the majority of TBI worldwide occurs in low- and middle-income countries. Further limitations stem from its observational design with pragmatic data collection. The broad inclusion criteria improve generalizability but potentially result in significant disparities between the CENTER-TBI and TRACK-TBI cohorts.^22–24^ CENTER-TBI patients were generally older and suffered more severe TBI compared to TRACK-TBI patients. These factors potentially delineate the apparent differences in mortality and unfavorable outcome between both cohorts. Care should be taken comparing the model performance between the CENTER-TBI and TRACK-TBI models. Discrepancies in case-mix, however, do not preclude comparative analysis of GCS versus GCS-P or GCS and PRS separately, and may even increase generalizability.

We cannot exclude the possibility that some imputation bias may have occurred. In CENTER-TBI a derived baseline GCS score was used with imputation of missing GCS scores according to IMPACT methodology. In TRACK-TBI the GCS scores were always calculated using the GCS ED admission scores and only imputed if the motor score was available.^14^

## Conclusions

GCS-P has a stronger association with outcome after TBI than the GCS alone, and provides a single integrated score. However, this comes at a loss of some information, and for prognostic models, inclusion of GCS and pupillary reactivity as separate scores is preferable to the use of a GCS-P summary score.

## Data Availability

All data produced in the present study are available upon reasonable request to the authors.

## Acknowledgements

We would like to gratefully thank The Clinical Assessment Work Group of the NIH-NINDS on classification and nomenclature of TBI and the CENTER-TBI and TRACK-TBI participants and investigators listed below.

## The Clinical Assessment Work Group of the NIH-NINDS

### Workgroup Leads

Adam R Ferguson (University of California San Francisco)

David K Menon (University of Cambridge)

Noah D Silverberg (University of British Columbia)

### Workgroup Members

Thomas J Bayuk (HCOS/NICoE/WRNMMC)

Matt Breiding (Centers for Disease Control and Prevention)

David L Brody (Uniformed Services University of the Health Sciences)

Todd A Cesar (TBICoE DHA)

Scott A Cota (TBICoE DHA)

Ari Ercole (University of Cambridge)

Anthony Figaji (University of Cape Town)

Guoyi Gao (Capital Medical University, Beijing)

Christopher Giza (University of California Los Angeles)

Fiona Lecky (University of Sheffield)

Rebekah Mannix (Boston Children’s Hospital)

Kasey Moritz (Combat Casualty Care Research Program)

Claudia S Robertson (Baylor College of Medicine)

John Yue (University of California San Francisco)

### Affiliate Members

Shubhayu Bhattacharyay (University of Cambridge)

Carl Marincowitz (University of Sheffield)

Ana Mikolic (University of British Columbia)

Abel Torres-Espin (University of Waterloo)

Spyridoula Tsetsou (Baylor College of Medicine)

## CENTER-TBI participants and investigators (to be indexed as “Collaborators” in PubMed)

Cecilia Åkerlund^1^, Krisztina Amrein^2^, Nada Andelic^3^, Lasse Andreassen^4^, Audny Anke^5^, Anna Antoni^6^, Gérard Audibert^7^, Philippe Azouvi^8^, Maria Luisa Azzolini^9^, Ronald Bartels^10^, Pál Barzó^11^, Romuald Beauvais^12^, Ronny Beer^13^, Bo-Michael Bellander^14^, Antonio Belli^15^, Habib Benali^16^, Maurizio Berardino^17^, Luigi Beretta^9^, Morten Blaabjerg^18^, Peter Bragge^19^, Alexandra Brazinova^20^, Vibeke Brinck^21^, Joanne Brooker^22^, Camilla Brorsson^23^, Andras Buki^24^, Monika Bullinger^25^, Manuel Cabeleira^26^, Alessio Caccioppola^27^, Emiliana Calappi ^27^, Maria Rosa Calvi^9^, Peter Cameron^28^, Guillermo Carbayo Lozano^29^, Marco Carbonara^27^, Simona Cavallo^17^, Giorgio Chevallard^30^, Arturo Chieregato^30^, Giuseppe Citerio^31, 32^, Hans Clusmann^33^, Mark Coburn^34^, Jonathan Coles^35^, Jamie D. Cooper^36^, Marta Correia^37^, Amra Čović ^38^, Nicola Curry^39^, Endre Czeiter^24^, Marek Czosnyka^26^, Claire Dahyot-Fizelier^40^, Paul Dark^41^, Helen Dawes^42^, Véronique De Keyser^43^, Vincent Degos^16^, Francesco Della Corte^44^, Hugo den Boogert^10^, Bart Depreitere^45^, Đula Đilvesi ^46^, Abhishek Dixit^47^, Emma Donoghue^22^, Jens Dreier^48^, Guy-Loup Dulière^49^, Ari Ercole^47^, Patrick Esser^42^, Erzsébet Ezer^50^, Martin Fabricius^51^, Valery L. Feigin^52^, Kelly Foks^53^, Shirin Frisvold^54^, Alex Furmanov^55^, Pablo Gagliardo^56^, Damien Galanaud^16^, Dashiell Gantner^28^, Guoyi Gao^57^, Pradeep George^58^, Alexandre Ghuysen^59^, Lelde Giga^60^, Ben Glocker^61^, Jagoš Golubovic^46^, Pedro A. Gomez ^62^, Johannes Gratz^63^, Benjamin Gravesteijn^64^, Francesca Grossi^44^, Russell L. Gruen^65^, Deepak Gupta^66^, Juanita A. Haagsma^64^, Iain Haitsma^67^, Raimund Helbok^13^, Eirik Helseth^68^, Lindsay Horton ^69^, Jilske Huijben^64^, Peter J. Hutchinson^70^, Bram Jacobs^71^, Stefan Jankowski^72^, Mike Jarrett^21^, Ji-yao Jiang^58^, Faye Johnson^73^, Kelly Jones^52^, Mladen Karan^46^, Angelos G. Kolias^70^, Erwin Kompanje^74^, Daniel Kondziella^51^, Evgenios Kornaropoulos^47^, Lars-Owe Koskinen^75^, Noémi Kovács^76^, Ana Kowark^77^, Alfonso Lagares^62^, Linda Lanyon^58^, Steven Laureys^78^, Fiona Lecky^79, 80^, Didier Ledoux^78^, Rolf Lefering^81^, Valerie Legrand^82^, Aurelie Lejeune^83^, Leon Levi^84^, Roger Lightfoot^85^, Hester Lingsma^64^, Andrew I.R. Maas^43^, Ana M. Castaño-León^62^, Marc Maegele^86^, Marek Majdan^20^, Alex Manara^87^, Geoffrey Manley^88^, Costanza Martino^89^, Hugues Maréchal^49^, Julia Mattern^90^, Catherine McMahon^91^, Béla Melegh^92^, David Menon^47^, Tomas Menovsky^43^, Ana Mikolic^64^, Benoit Misset^78^, Visakh Muraleedharan^58^, Lynnette Murray^28^, Ancuta Negru^93^, David Nelson^1^, Virginia Newcombe^47^, Daan Nieboer^64^, József Nyirádi^2^, Otesile Olubukola^79^, Matej Oresic^94^, Fabrizio Ortolano^27^, Aarno Palotie^95, 96, 97^, Paul M. Parizel^98^, Jean-François Payen^99^, Natascha Perera^12^, Vincent Perlbarg^16^, Paolo Persona^100^, Wilco Peul^101^, Anna Piippo-Karjalainen^102^, Matti Pirinen^95^, Dana Pisica^64^, Horia Ples^93^, Suzanne Polinder^64^, Inigo Pomposo^29^, Jussi P. Posti ^103^, Louis Puybasset^104^, Andreea Radoi ^105^, Arminas Ragauskas^106^, Rahul Raj^102^, Malinka Rambadagalla^107^, Isabel Retel Helmrich^64^, Jonathan Rhodes^108^, Sylvia Richardson^109^, Sophie Richter^47^, Samuli Ripatti^95^, Saulius Rocka^106^, Cecilie Roe^110^, Olav Roise^111,112^, Jonathan Rosand^113^, Jeffrey V. Rosenfeld^114^, Christina Rosenlund^115^, Guy Rosenthal^55^, Rolf Rossaint^77^, Sandra Rossi^100^, Daniel Rueckert^61^ Martin Rusnák^116^, Juan Sahuquillo^105^, Oliver Sakowitz^90, 117^, Renan Sanchez-Porras^117^, Janos Sandor^118^, Nadine Schäfer^81^, Silke Schmidt^119^, Herbert Schoechl^120^, Guus Schoonman^121^, Rico Frederik Schou^122^, Elisabeth Schwendenwein^6^, Charlie Sewalt^64^, Ranjit D. Singh^101^, Toril Skandsen^123, 124^, Peter Smielewski^26^, Abayomi Sorinola^125^, Emmanuel Stamatakis^47^, Simon Stanworth^39^, Robert Stevens^126^, William Stewart^127^, Ewout W. Steyerberg^64, 128^, Nino Stocchetti^129^, Nina Sundström^130^, Riikka Takala^131^, Viktória Tamás^125^, Tomas Tamosuitis^132^, Mark Steven Taylor^20^, Aurore Thibaut^78^, Braden Te Ao^52^, Olli Tenovuo^103^, Alice Theadom^52^, Matt Thomas^87^, Dick Tibboel^133^, Marjolein Timmers^74^, Christos Tolias^134^, Tony Trapani^28^, Cristina Maria Tudora^93^, Andreas Unterberg^90^, Peter Vajkoczy ^135^, Shirley Vallance^28^, Egils Valeinis^60^, Zoltán Vámos^50^, Mathieu van der Jagt^136^, Gregory Van der Steen^43^, Joukje van der Naalt^71^, Jeroen T.J.M. van Dijck ^101^, Inge A. M. van Erp^101^, Thomas A. van Essen^101^, Wim Van Hecke^137^, Caroline van Heugten^138^, Dominique Van Praag^139^, Ernest van Veen^64^, Thijs Van de Vyvere^137^, Roel P. J. van Wijk^101^, Alessia Vargiolu^32^, Emmanuel Vega^83^, Kimberley Velt^64^, Jan Verheyden^137^, Paul M. Vespa^140^, Anne Vik^123, 141^, Rimantas Vilcinis^132^, Victor Volovici^67^, Nicole von Steinbüchel^38^, Daphne Voormolen^64^, Rick J.G. Vreeburg^101^, Petar Vulekovic^46^, Kevin K.W. Wang^142^, Daniel Whitehouse^47^, Eveline Wiegers^64^, Guy Williams^47^, Lindsay Wilson^69^, Stefan Winzeck^47^, Stefan Wolf^143^, Zhihui Yang^113^, Peter Ylén^144^, Alexander Younsi^90^, Frederick A. Zeiler^47,145^, Veronika Zelinkova^20^, Agate Ziverte^60^, Tommaso Zoerle^27^

^1^ Department of Physiology and Pharmacology, Section of Perioperative Medicine and Intensive Care, Karolinska Institutet, Stockholm, Sweden

^2^ János Szentágothai Research Centre, University of Pécs, Pécs, Hungary

^3^ Division of Clinical Neuroscience, Department of Physical Medicine and Rehabilitation, Oslo University Hospital and University of Oslo, Oslo, Norway

^4^ Department of Neurosurgery, University Hospital Northern Norway, Tromso, Norway

^5^ Department of Physical Medicine and Rehabilitation, University Hospital Northern Norway, Tromso, Norway

^6^ Trauma Surgery, Medical University Vienna, Vienna, Austria

^7^ Department of Anesthesiology & Intensive Care, University Hospital Nancy, Nancy, France

^8^ Raymond Poincare hospital, Assistance Publique – Hopitaux de Paris, Paris, France

^9^ Department of Anesthesiology & Intensive Care, S Raffaele University Hospital, Milan, Italy

^10^ Department of Neurosurgery, Radboud University Medical Center, Nijmegen, The Netherlands

^11^ Department of Neurosurgery, University of Szeged, Szeged, Hungary

^12^ International Projects Management, ARTTIC, Munchen, Germany

^13^ Department of Neurology, Neurological Intensive Care Unit, Medical University of Innsbruck, Innsbruck, Austria

^14^ Department of Neurosurgery & Anesthesia & intensive care medicine, Karolinska University Hospital, Stockholm, Sweden

^15^ NIHR Surgical Reconstruction and Microbiology Research Centre, Birmingham, UK

^16^ Anesthesie-Réanimation, Assistance Publique – Hopitaux de Paris, Paris, France

^17^ Department of Anesthesia & ICU, AOU Città della Salute e della Scienza di Torino - Orthopedic and Trauma Center, Torino, Italy

^18^ Department of Neurology, Odense University Hospital, Odense, Denmark

^19^ BehaviourWorks Australia, Monash Sustainability Institute, Monash University, Victoria, Australia

^20^ Department of Public Health, Faculty of Health Sciences and Social Work, Trnava University, Trnava, Slovakia

^21^ Quesgen Systems Inc., Burlingame, California, USA

^22^ Australian & New Zealand Intensive Care Research Centre, Department of Epidemiology and Preventive Medicine, School of Public Health and Preventive Medicine, Monash University, Melbourne, Australia

^23^ Department of Surgery and Perioperative Science, Umeå University, Umeå, Sweden

^24^ Department of Neurosurgery, Medical School, University of Pécs, Hungary and Neurotrauma Research Group, János Szentágothai Research Centre, University of Pécs, Hungary

^25^ Department of Medical Psychology, Universitätsklinikum Hamburg-Eppendorf, Hamburg, Germany

^26^ Brain Physics Lab, Division of Neurosurgery, Dept of Clinical Neurosciences, University of Cambridge, Addenbrooke’s Hospital, Cambridge, UK

^27^ Neuro ICU, Fondazione IRCCS Cà Granda Ospedale Maggiore Policlinico, Milan, Italy

^28^ ANZIC Research Centre, Monash University, Department of Epidemiology and Preventive Medicine, Melbourne, Victoria, Australia

^29^ Department of Neurosurgery, Hospital of Cruces, Bilbao, Spain

^30^ NeuroIntensive Care, Niguarda Hospital, Milan, Italy

^31^ School of Medicine and Surgery, Università Milano Bicocca, Milano, Italy

^32^ NeuroIntensive Care Unit, Department Neuroscience, IRCCS Fondazione San Gerardo dei Tintori, Monza, Italy

^33^ Department of Neurosurgery, Medical Faculty RWTH Aachen University, Aachen, Germany

^34^ Department of Anesthesiology and Intensive Care Medicine, University Hospital Bonn, Bonn, Germany

^35^ Department of Anesthesia & Neurointensive Care, Cambridge University Hospital NHS Foundation Trust, Cambridge, UK

^36^ School of Public Health & PM, Monash University and The Alfred Hospital, Melbourne, Victoria, Australia

^37^ Radiology/MRI department, MRC Cognition and Brain Sciences Unit, Cambridge, UK

^38^ Institute of Medical Psychology and Medical Sociology, Universitätsmedizin Göttingen, Göttingen, Germany

^39^ Oxford University Hospitals NHS Trust, Oxford, UK

^40^ Intensive Care Unit, CHU Poitiers, Potiers, France

^41^ University of Manchester NIHR Biomedical Research Centre, Critical Care Directorate, Salford Royal Hospital NHS Foundation Trust, Salford, UK

^42^ Movement Science Group, Faculty of Health and Life Sciences, Oxford Brookes University, Oxford, UK

^43^ Department of Neurosurgery, Antwerp University Hospital and University of Antwerp, Edegem, Belgium

^44^ Department of Anesthesia & Intensive Care, Maggiore Della Carità Hospital, Novara, Italy

^45^ Department of Neurosurgery, University Hospitals Leuven, Leuven, Belgium

^46^ Department of Neurosurgery, Clinical centre of Vojvodina, Faculty of Medicine, University of Novi Sad, Novi Sad, Serbia

^47^ Division of Anaesthesia, University of Cambridge, Addenbrooke’s Hospital, Cambridge, UK

^48^ Center for Stroke Research Berlin, Charité – Universitätsmedizin Berlin, corporate member of Freie Universität Berlin, Humboldt-Universität zu Berlin, and Berlin Institute of Health, Berlin, Germany

^49^ Intensive Care Unit, CHR Citadelle, Liège, Belgium

^50^ Department of Anaesthesiology and Intensive Therapy, University of Pécs, Pécs, Hungary

^51^ Departments of Neurology, Clinical Neurophysiology and Neuroanesthesiology, Region Hovedstaden Rigshospitalet, Copenhagen, Denmark

^52^ National Institute for Stroke and Applied Neurosciences, Faculty of Health and Environmental Studies, Auckland University of Technology, Auckland, New Zealand

^53^ Department of Neurology, Erasmus MC, Rotterdam, the Netherlands

^54^ Department of Anesthesiology and Intensive care, University Hospital Northern Norway, Tromso, Norway

^55^ Department of Neurosurgery, Hadassah-hebrew University Medical center, Jerusalem, Israel

^56^ Fundación Instituto Valenciano de Neurorrehabilitación (FIVAN), Valencia, Spain

^57^ Department of Neurosurgery, Shanghai Renji hospital, Shanghai Jiaotong University/school of medicine, Shanghai, China

^58^ Karolinska Institutet, INCF International Neuroinformatics Coordinating Facility, Stockholm, Sweden

^59^ Emergency Department, CHU, Liège, Belgium

^60^ Neurosurgery clinic, Pauls Stradins Clinical University Hospital, Riga, Latvia

^61^ Department of Computing, Imperial College London, London, UK

^62^ Department of Neurosurgery, Hospital Universitario 12 de Octubre, Madrid, Spain

^63^ Department of Anesthesia, Critical Care and Pain Medicine, Medical University of Vienna, Austria

^64^ Department of Public Health, Erasmus Medical Center-University Medical Center, Rotterdam, The Netherlands

^65^ College of Health and Medicine, Australian National University, Canberra, Australia

^66^ Department of Neurosurgery, Neurosciences Centre & JPN Apex trauma centre, All India Institute of Medical Sciences, New Delhi-110029, India

^67^ Department of Neurosurgery, Erasmus MC, Rotterdam, the Netherlands

^68^ Department of Neurosurgery, Oslo University Hospital, Oslo, Norway

^69^ Division of Psychology, University of Stirling, Stirling, UK

^70^ Division of Neurosurgery, Department of Clinical Neurosciences, Addenbrooke’s Hospital & University of Cambridge, Cambridge, UK

^71^ Department of Neurology, University of Groningen, University Medical Center Groningen, Groningen, Netherlands

^72^ Neurointensive Care, Sheffield Teaching Hospitals NHS Foundation Trust, Sheffield, UK

^73^ Salford Royal Hospital NHS Foundation Trust Acute Research Delivery Team, Salford, UK

^74^ Department of Intensive Care and Department of Ethics and Philosophy of Medicine, Erasmus Medical Center, Rotterdam, The Netherlands

^75^ Department of Clinical Neuroscience, Neurosurgery, Umeå University, Umeå, Sweden

^76^ Hungarian Brain Research Program - Grant No. KTIA_13_NAP-A-II/8, University of Pécs, Pécs, Hungary

^77^ Department of Anaesthesiology, University Hospital of Aachen, Aachen, Germany

^78^ Cyclotron Research Center, University of Liège, Liège, Belgium

^79^ Centre for Urgent and Emergency Care Research (CURE), Health Services Research Section, School of Health and Related Research (ScHARR), University of Sheffield, Sheffield, UK

^80^ Emergency Department, Salford Royal Hospital, Salford UK

^81^ Institute of Research in Operative Medicine (IFOM), Witten/Herdecke University, Cologne, Germany

^82^ VP Global Project Management CNS, ICON, Paris, France

^83^ Department of Anesthesiology-Intensive Care, Lille University Hospital, Lille, France

^84^ Department of Neurosurgery, Rambam Medical Center, Haifa, Israel

^85^ Department of Anesthesiology & Intensive Care, University Hospitals Southhampton NHS Trust, Southhampton, UK

^86^ Cologne-Merheim Medical Center (CMMC), Department of Traumatology, Orthopedic Surgery and Sportmedicine, Witten/Herdecke University, Cologne, Germany

^87^ Intensive Care Unit, Southmead Hospital, Bristol, Bristol, UK

^88^ Department of Neurological Surgery, University of California, San Francisco, California, USA

^89^ Department of Anesthesia & Intensive Care,M. Bufalini Hospital, Cesena, Italy

^90^ Department of Neurosurgery, University Hospital Heidelberg, Heidelberg, Germany

^91^ Department of Neurosurgery, The Walton centre NHS Foundation Trust, Liverpool, UK

^92^ Department of Medical Genetics, University of Pécs, Pécs, Hungary

^93^ Department of Neurosurgery, Emergency County Hospital Timisoara, Timisoara, Romania

^94^ School of Medical Sciences, Örebro University, Örebro, Sweden

^95^ Institute for Molecular Medicine Finland, University of Helsinki, Helsinki, Finland

^96^ Analytic and Translational Genetics Unit, Department of Medicine; Psychiatric & Neurodevelopmental Genetics Unit, Department of Psychiatry; Department of Neurology, Massachusetts General Hospital, Boston, MA, USA

^97^ Program in Medical and Population Genetics; The Stanley Center for Psychiatric Research, The Broad Institute of MIT and Harvard, Cambridge, MA, USA

^98^ Department of Radiology, University of Antwerp, Edegem, Belgium

^99^ Department of Anesthesiology & Intensive Care, University Hospital of Grenoble, Grenoble, France

^100^ Department of Anesthesia & Intensive Care, Azienda Ospedaliera Università di Padova, Padova, Italy

^101^ Dept. of Neurosurgery, Leiden University Medical Center, Leiden, The Netherlands and Dept. of Neurosurgery, Medical Center Haaglanden, The Hague, The Netherlands

^102^ Department of Neurosurgery, Helsinki University Central Hospital

^103^ Division of Clinical Neurosciences, Department of Neurosurgery and Turku Brain Injury Centre, Turku University Hospital and University of Turku, Turku, Finland

^104^ Department of Anesthesiology and Critical Care, Pitié -Salpêtrière Teaching Hospital, Assistance Publique, Hôpitaux de Paris and University Pierre et Marie Curie, Paris, France

^105^ Neurotraumatology and Neurosurgery Research Unit (UNINN), Vall d’Hebron Research Institute, Barcelona, Spain

^106^ Department of Neurosurgery, Kaunas University of technology and Vilnius University, Vilnius, Lithuania

^107^ Department of Neurosurgery, Rezekne Hospital, Latvia

^108^ Department of Anaesthesia, Critical Care & Pain Medicine NHS Lothian & University of Edinburg, Edinburgh, UK

^109^ Director, MRC Biostatistics Unit, Cambridge Institute of Public Health, Cambridge, UK

^110^ Department of Physical Medicine and Rehabilitation, Oslo University Hospital/University of Oslo, Oslo, Norway

^111^ Division of Orthopedics, Oslo University Hospital, Oslo, Norway

^112^ Institue of Clinical Medicine, Faculty of Medicine, University of Oslo, Oslo, Norway

^113^ Broad Institute, Cambridge MA Harvard Medical School, Boston MA, Massachusetts General Hospital, Boston MA, USA

^114^ National Trauma Research Institute, The Alfred Hospital, Monash University, Melbourne, Victoria, Australia

^115^ Department of Neurosurgery, Odense University Hospital, Odense, Denmark

^116^ International Neurotrauma Research Organisation, Vienna, Austria

^117^ Klinik für Neurochirurgie, Klinikum Ludwigsburg, Ludwigsburg, Germany

^118^ Division of Biostatistics and Epidemiology, Department of Preventive Medicine, University of Debrecen, Debrecen, Hungary

^119^ Department Health and Prevention, University Greifswald, Greifswald, Germany

^120^ Department of Anaesthesiology and Intensive Care, AUVA Trauma Hospital, Salzburg, Austria

^121^ Department of Neurology, Elisabeth-TweeSteden Ziekenhuis, Tilburg, the Netherlands

^122^ Department of Neuroanesthesia and Neurointensive Care, Odense University Hospital, Odense, Denmark

^123^ Department of Neuromedicine and Movement Science, Norwegian University of Science and Technology, NTNU, Trondheim, Norway

^124^ Department of Physical Medicine and Rehabilitation, St.Olavs Hospital, Trondheim University Hospital, Trondheim, Norway

^125^ Department of Neurosurgery, University of Pécs, Pécs, Hungary

^126^ Division of Neuroscience Critical Care, John Hopkins University School of Medicine, Baltimore, USA

^127^ Department of Neuropathology, Queen Elizabeth University Hospital and University of Glasgow, Glasgow, UK

^128^ Dept. of Department of Biomedical Data Sciences, Leiden University Medical Center, Leiden, The Netherlands

^129^ Department of Pathophysiology and Transplantation, Milan University, and Neuroscience ICU, Fondazione IRCCS Cà Granda Ospedale Maggiore Policlinico, Milano, Italy

^130^ Department of Radiation Sciences, Biomedical Engineering, Umeå University, Umeå, Sweden

^131^ Perioperative Services, Intensive Care Medicine and Pain Management, Turku University Hospital and University of Turku, Turku, Finland

^132^ Department of Neurosurgery, Kaunas University of Health Sciences, Kaunas, Lithuania

^133^ Intensive Care and Department of Pediatric Surgery, Erasmus Medical Center, Sophia Children’s Hospital, Rotterdam, The Netherlands

^134^ Department of Neurosurgery, Kings college London, London, UK

^135^ Neurologie, Neurochirurgie und Psychiatrie, Charité – Universitätsmedizin Berlin, Berlin, Germany

^136^ Department of Intensive Care Adults, Erasmus MC– University Medical Center Rotterdam, Rotterdam, the Netherlands

^137^ icoMetrix NV, Leuven, Belgium

^138^ Movement Science Group, Faculty of Health and Life Sciences, Oxford Brookes University, Oxford, UK

^139^ Psychology Department, Antwerp University Hospital, Edegem, Belgium

^140^ Director of Neurocritical Care, University of California, Los Angeles, USA

^141^ Department of Neurosurgery, St.Olavs Hospital, Trondheim University Hospital, Trondheim, Norway

^142^ Department of Emergency Medicine, University of Florida, Gainesville, Florida, USA

^143^ Department of Neurosurgery, Charité – Universitätsmedizin Berlin, corporate member of Freie Universität Berlin, Humboldt-Universität zu Berlin, and Berlin Institute of Health, Berlin, Germany

^144^ VTT Technical Research Centre, Tampere, Finland

^145^ Section of Neurosurgery, Department of Surgery, Rady Faculty of Health Sciences, University of Manitoba, Winnipeg, MB, Canada

## TRACK-TBI Investigators (to be indexed as “Collaborators” in PubMed)

Neeraj Badjatia, MD, MS (Department of Neurology, University of Maryland, Baltimore, Maryland, US); Jason Barber, MS (Department of Neurological Surgery, University of Washington, Seattle, Washington, US); Yelena G. Bodien, PhD (Department of Neurology, Harvard Medical School, Boston, Massachusetts, US); Brian Fabian, MPA (Department of Neurological Surgery, University of California, San Francisco, San Francisco, California, US); Adam R. Ferguson, PhD (Department of Neurological Surgery, University of California, San Francisco, San Francisco, California, US); Brandon Foreman, MD (Department of Neurology, University of Cincinnati, Cincinnati, Ohio, US); Raquel C. Gardner, MD (Department of Neurology, University of California, San Francisco, San Francisco, California, US); Shankar Gopinath, MD (Department of Neurological Surgery, Baylor College of Medicine, Houston, Texas, US); Ramesh Grandhi, MD, MS (Department of Neurological Surgery, University of Utah Medical Center; Salt Lake City, Utah, US); J. Russell Huie, PhD (Department of Neurological Surgery, University of California, San Francisco, San Francisco, California, US); C. Dirk Keene, MD, PhD (Department of Laboratory Medicine and Pathology, University of Washington, Seattle, Washington, US); Hester F. Lingsma, PhD (Department of Public Health, Erasmus Medical Center, Rotterdam, The Netherlands); Christine L. Mac Donald, PhD (Department of Neurological Surgery, University of Washington, Seattle, Washington, US); Amy J. Markowitz, JD (Department of Neurological Surgery, University of California, San Francisco, San Francisco, California, US); Randall Merchant, PhD (Department of Anatomy, Virginia Commonwealth University, Richmond, Virginia, US); Laura B. Ngwenya, MD, PhD (Department of Neurological Surgery, University of Cincinnati, Cincinnati, Ohio, US); Richard B. Rodgers, MD (Department of Neurosurgery, Goodman Campbell Brain and Spine, Indianapolis, Indiana, US); Andrea L. C. Schneider, MD, PhD (Department of Neurology, University of Pennsylvania, Philadelphia, PA, US); David M. Schnyer, PhD (Department of Psychology, University of Texas at Austin, Austin, Texas, US); Sabrina R. Taylor, PhD (Department of Neurological Surgery, University of California, San Francisco, San Francisco, California, US); Nancy R. Temkin, PhD (Department of Neurological Surgery, University of Washington, Seattle, Washington, US); Abel Torres-Espin, PhD (Department of Neurological Surgery, University of California, San Francisco, San Francisco, California, US); Mary J. Vassar, RN, MS (Department of Neurological Surgery, University of California, San Francisco, San Francisco, California, US); Kevin K. W. Wang, PhD (Department of Neurobiology, Morehouse School of Medicine, Atlanta, Georgia, US); Justin C. Wong, BS (Department of Neurological Surgery, University of California, San Francisco, San Francisco, California, US); Ross D. Zafonte, DO (Department of Rehabilitation Medicine, Harvard Medical School, Boston, Massachusetts, US)

