## Supplementary figures and images for "Validation of the GCS-Pupil scale in Traumatic Brain Injury Incremental prognostic performance of pupillary reactivity with GCS in the prospective observational cohorts CENTER-TBI and TRACK-TBI"

### Supplemental Figure 1

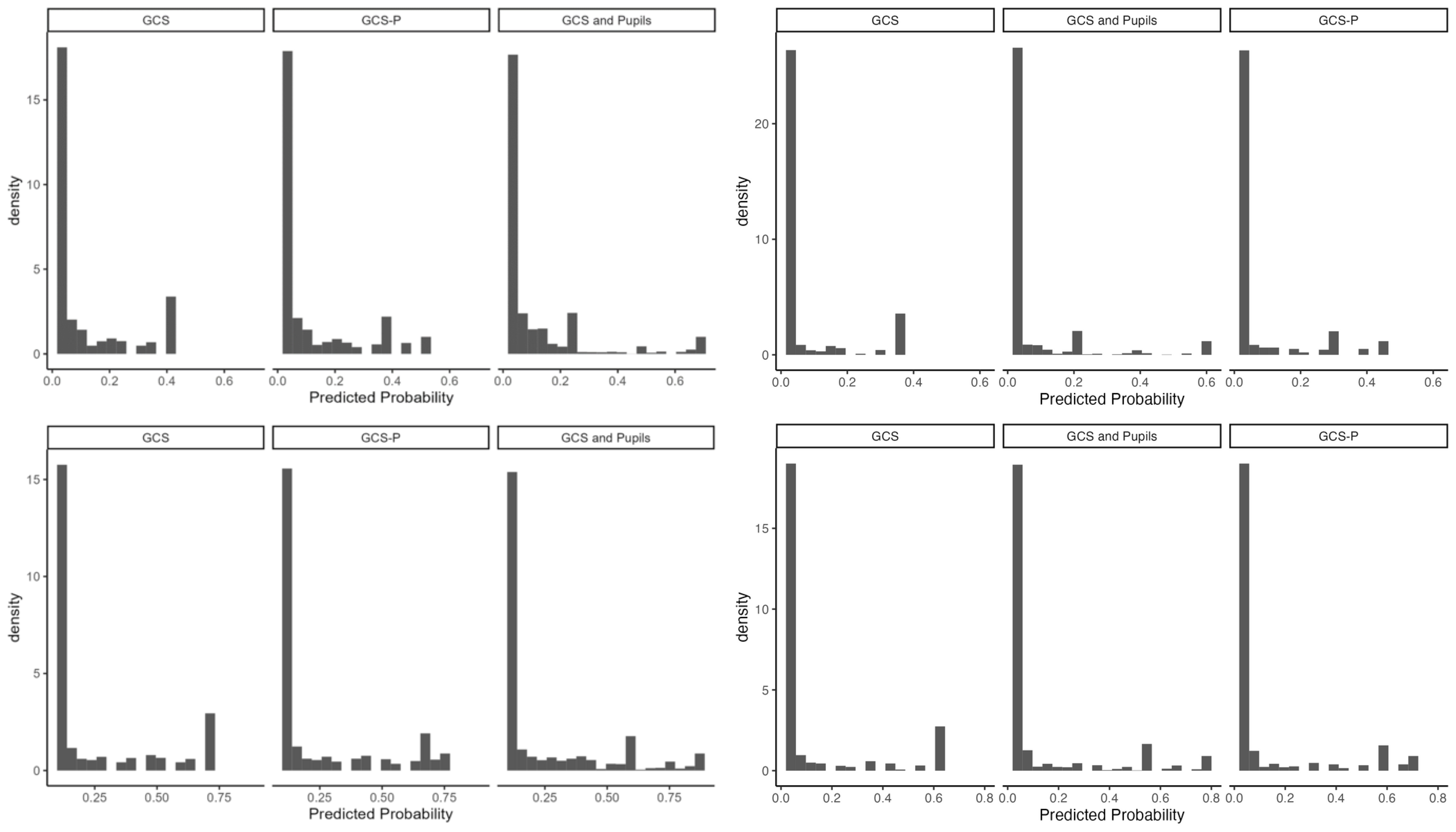

### Supplemental Figure 2

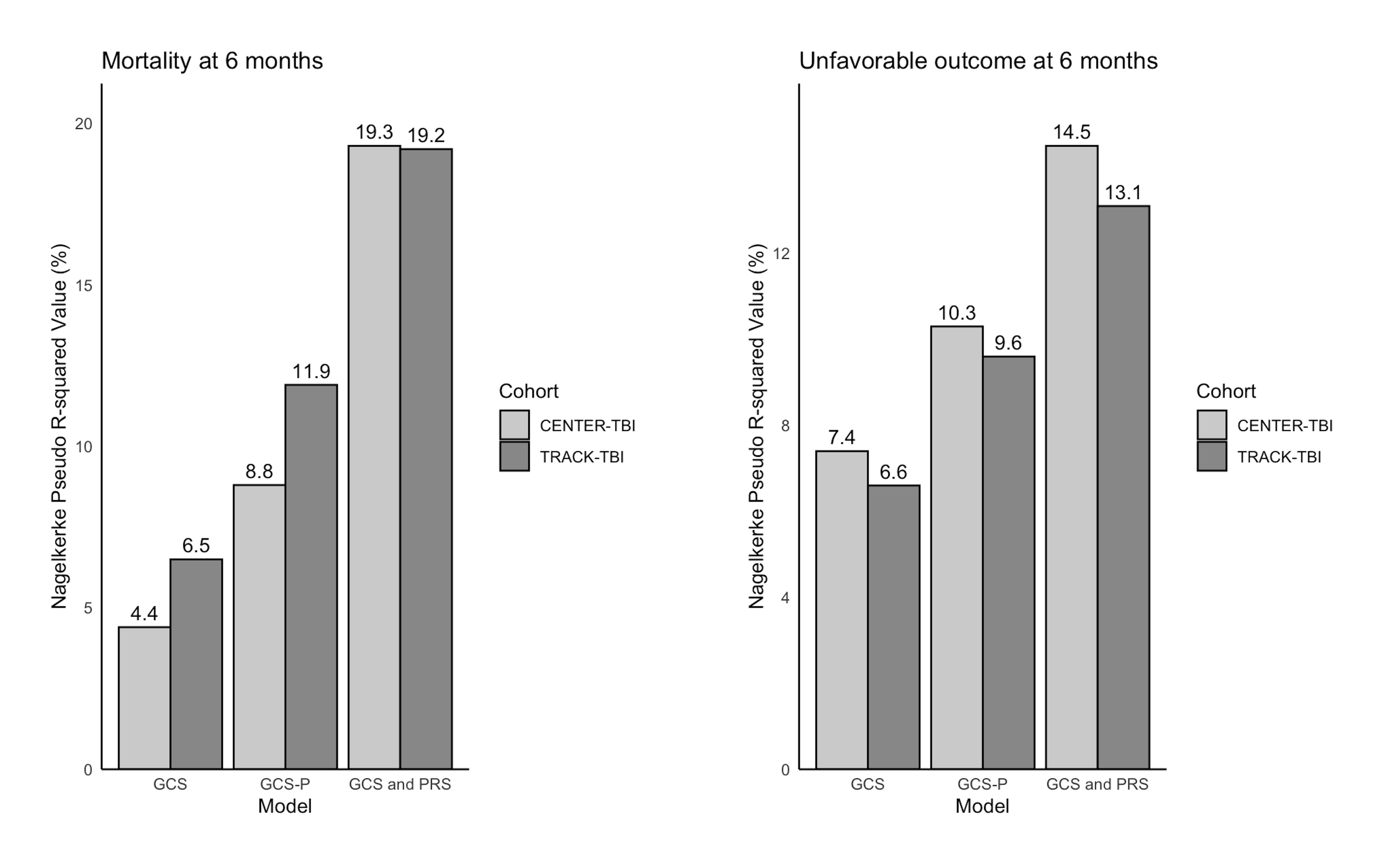
