## Supplemental Table 1 for "Validation of the GCS-Pupil scale in Traumatic Brain Injury Incremental prognostic performance of pupillary reactivity with GCS in the prospective observational cohorts CENTER-TBI and TRACK-TBI"

**Supplemental Table 1 | Mortality and unfavorable outcome distribution by GCS and pupil reactivity.**

*CENTER-TBI*

|  | **Both reacted** | | | **One reacted** | | | **None reacted** | | |  |
| --- | --- | --- | --- | --- | --- | --- | --- | --- | --- | --- |
| **GCS Score** | **No** | **% Dead at 6 Mo** | **% Unfav at 6 Mo** | **No** | **% Dead at 6 Mo** | **% Unfav at 6 Mo** | **No** | **% Dead at 6 Mo** | **% Unfav at 6 Mo** | **Total No. Patients** |
| 3 | 242 | 22 | 51 | 47 | 43 | 70 | 122 | 69 | 84 | 411 |
| 4 | 41 | 15 | 68 | 11 | 36 | 82 | 31 | 74 | 94 | 83 |
| 5 | 35 | 17 | 60 | 10 | 30 | 60 | 14 | 43 | 64 | 59 |
| 6 | 69 | 22 | 51 | 5 | 60 | 80 | 17 | 59 | 76 | 91 |
| 7 | 98 | 11 | 36 | 5 | 0 | 40 | 8 | 38 | 50 | 111 |
| 8 | 76 | 13 | 39 | 7 | 29 | 57 | 7 | 43 | 57 | 90 |
| 9 | 54 | 24 | 39 | 4 | 25 | 50 | 1 | 100 | 100 | 59 |
| 10 | 89 | 21 | 43 | 4 | 25 | 50 | 5 | 80 | 80 | 98 |
| 11 | 66 | 20 | 36 | 3 | 33 | 67 | 6 | 33 | 67 | 75 |
| 12 | 74 | 14 | 26 | 3 | 33 | 67 | 7 | 43 | 57 | 84 |
| 13 | 151 | 10 | 23 | 5 | 20 | 20 | 6 | 50 | 83 | 162 |
| 14 | 405 | 6 | 16 | 8 | 13 | 38 | 6 | 17 | 33 | 419 |
| 15 | 1741 | 2 | 8 | 25 | 0 | 20 | 13 | 15 | 15 | 1779 |
| Total | 3141 | 15 | 38 | 137 | 27 | 54 | 243 | 51 | 66 |  |

*Abbreviations*: CENTER-TBI, Collaborative European NeuroTrauma Effectiveness Research in Traumatic Brain Injury; GCS, Glasgow Coma Scale; Mo, month; No, number; Unfav, unfavorable outcome

*TRACK-TBI*

|  | **Both reacted** | | | **One reacted** | | | **None reacted** | | |  |
| --- | --- | --- | --- | --- | --- | --- | --- | --- | --- | --- |
| **GCS Score** | **No** | **% Dead at 6 Mo** | **% Unfav at 6 Mo** | **No** | **% Dead at 6 Mo** | **% Unfav at 6 Mo** | **No** | **% Dead at 6 Mo** | **% Unfav at 6 Mo** | **Total No. Patients** |
| 3 | 114 | 15 | 35 | 18 | 44 | 72 | 68 | 40 | 53 | 200 |
| 4 | 15 | 7 | 40 | 1 | 0 | 100 | 7 | 14 | 43 | 23 |
| 5 | 4 | 0 | 25 | 1 | 100 | 100 | 2 | 0 | 50 | 7 |
| 6 | 23 | 4 | 17 | 3 | 0 | 0 | 6 | 67 | 83 | 32 |
| 7 | 27 | 15 | 33 | 3 | 100 | 100 | 7 | 29 | 43 | 37 |
| 8 | 21 | 5 | 14 | 1 | 0 | 0 | 1 | 100 | 100 | 23 |
| 9 | 23 | 0 | 22 | 3 | 0 | 0 | 4 | 50 | 75 | 30 |
| 10 | 33 | 18 | 30 | 0 | 0 | 0 | 1 | 0 | 0 | 34 |
| 11 | 21 | 0 | 29 | 0 | 0 | 0 | 0 | 0 | 0 | 21 |
| 12 | 29 | 0 | 0 | 0 | 0 | 0 | 0 | 0 | 0 | 29 |
| 13 | 80 | 1 | 4 | 2 | 50 | 50 | 1 | 0 | 0 | 83 |
| 14 | 338 | 2 | 5 | 2 | 0 | 0 | 0 | 0 | 0 | 340 |
| 15 | 1318 | 1 | 2 | 7 | 0 | 0 | 0 | 0 | 0 | 1325 |
| Total | 2046 | 2 | 6 | 41 | 32 | 46 | 97 | 38 | 54 |  |

*Abbreviations*: GCS, Glasgow Coma Scale; Mo, month; No, number; TRACK-TBI, Transforming Research and Clinical Knowledge in Traumatic Brain Injury; Unfav, unfavorable outcome
