## Supplemental Table 2 for "Validation of the GCS-Pupil scale in Traumatic Brain Injury Incremental prognostic performance of pupillary reactivity with GCS in the prospective observational cohorts CENTER-TBI and TRACK-TBI"

**Supplemental Table 2 | Mortality and unfavorable outcome distribution by GCS-components and PRS**

|  | **CENTER-TBI** | | | **TRACK-TBI** | | |
| --- | --- | --- | --- | --- | --- | --- |
| **GCS component combination^a^** | **No. Patients** | **Dead at 6 Mo (%)^b^** | **Unfav at 6 Mo (%)^b^** | **No. Patients** | **Dead at 6 Mo (%)^b^** | **Unfav at 6 Mo (%)^b^** |
| E1 V1 M1 P0 | 340 | 72 (21) | 168 (49) | 41 | 9 (22) | 19 (46) |
| E1 V1 M2 P1 | 4 | 2 (50) | 4 (100) | 1 | 0 (0) | 1 (100) |
| E1 V1 M3 P2 | 10 | 6 (60) | 8 (80) | 1 | 0 (0) | 1 (100) |

*Abbreviations*: CENTER-TBI, Collaborative European NeuroTrauma Effectiveness Research in Traumatic Brain Injury; GCS-P, Glasgow Coma Scale – Pupils; Mo, month; No, number; TRACK-TBI, Transforming Research and Clinical Knowledge in Traumatic Brain Injury

^a^No patients were included with other combinations equaling a GCS-P score of 3.

^b^Pooled comparison of both cohorts yielded statistically significant differences in mortality and unfavorable outcome within the different GCS-P equals 3 combinations (mortality p-value <0.01 and chi-square statistic: 9.2, unfavorable outcome p-value <0.01) and chi-square statistic: 10).
