## Supplemental Table 3 for "Validation of the GCS-Pupil scale in Traumatic Brain Injury Incremental prognostic performance of pupillary reactivity with GCS in the prospective observational cohorts CENTER-TBI and TRACK-TBI"

**Supplemental Table 3 | Mortality and unfavorable outcome distribution by GCS-P**

|  | **CENTER-TBI** | | | **TRACK-TBI** | | |
| --- | --- | --- | --- | --- | --- | --- |
| **GCS-P Score** | **No. Patients** | **% Dead at 6 Mo** | **% Unfav at 6 Mo** | **No. Patients** | **% Dead at 6 Mo** | **% Unfav at 6 Mo** |
| 1 | 122 | 69 | 84 | 51 | 53 | 71 |
| 2 | 78 | 55 | 79 | 22 | 41 | 73 |
| 3 | 267 | 24 | 53 | 88 | 19 | 48 |
| 4 | 68 | 28 | 69 | 19 | 32 | 63 |
| 5 | 48 | 25 | 60 | 9 | 22 | 44 |
| 6 | 81 | 22 | 51 | 22 | 23 | 36 |
| 7 | 106 | 13 | 38 | 27 | 22 | 44 |
| 8 | 85 | 18 | 42 | 15 | 7 | 20 |
| 9 | 64 | 25 | 42 | 12 | 0 | 42 |
| 10 | 99 | 23 | 44 | 24 | 25 | 42 |
| 11 | 75 | 23 | 41 | 13 | 0 | 46 |
| 12 | 85 | 14 | 26 | 18 | 6 | 6 |
| 13 | 172 | 10 | 23 | 51 | 2 | 6 |
| 14 | 430 | 5 | 17 | 231 | 3 | 7 |
| 15 | 1741 | 2 | 8 | 837 | 1 | 3 |
| Total | 3521 | 24 | 45 | 1439 | 7 | 14 |

*Abbreviations*: CENTER-TBI, Collaborative European NeuroTrauma Effectiveness Research in Traumatic Brain Injury; GCS-P, Glasgow Coma Scale – Pupils; Mo, month; No, number; TRACK-TBI, Transforming Research and Clinical Knowledge in Traumatic Brain Injury
