## Supplemental Table 4 for "Validation of the GCS-Pupil scale in Traumatic Brain Injury Incremental prognostic performance of pupillary reactivity with GCS in the prospective observational cohorts CENTER-TBI and TRACK-TBI"

**Supplemental Table 4 | Overall predictive performance of different regression models in subgroups**

| **CENTER-TBI** | | | **TRACK-TBI** | |
| --- | --- | --- | --- | --- |
| **Model Specifications** | **Nagelkerke’s R^2^ (%) for Mortality** | **Nagelkerke’s R^2^ (%) for Unfavorable outcome** | **Nagelkerke’s R^2^ (%) for Mortality** | **Nagelkerke’s R^2^ (%) for Unfavorable outcome** |
| ***GCS < 13*** |  |  |  |  |
| GCS | 4.4 | 7.4 | 6.5 | 6.6 |
| GCS-P | 8.8 | 10 | 12 | 9.6 |
| GCS + PRS | 19 | 15 | 19 | 13 |
| ***Age < 45*** |  |  |  |  |
| GCS | 37 | 41 | 34 | 44 |
| GCS-P | 43 | 43 | 38 | 47 |
| GCS + PRS | 47 | 44 | 42 | 48 |
| ***Age 45-64*** |  |  |  |  |
| GCS | 25 | 31 | 31 | 41 |
| GCS-P | 28 | 32 | 33 | 42 |
| GCS + PRS | 31 | 32 | 36 | 42 |
| ***Age ≥ 65*** |  |  |  |  |
| GCS | 34 | 35 | 52 | 52 |
| GCS-P | 36 | 36 | 54 | 52 |
| GCS + PRS | 38 | 37 | 55 | 52 |

*Abbreviations*: CENTER-TBI, Collaborative European NeuroTrauma Effectiveness Research in Traumatic Brain Injury; GCS, Glasgow Coma Scale; GCS-P, Glasgow Coma Scale – Pupils; IQR, interquartile range; No, number; PRS, pupils reactivity score; TRACK-TBI, Transforming Research and Clinical Knowledge in Traumatic Brain Injury
